# Type of refugee accommodation and health of residents: cross-sectional cluster analysis

**DOI:** 10.1101/2022.12.11.22283314

**Authors:** Amir Mohsenpour, Verena Dudek, Kayvan Bozorgmehr, Louise Biddle, Oliver Razum, Odile Sauzet

## Abstract

Few studies have assessed whether refugees’ health is associated with accommodation characteristics. We aimed to devise a typology of refugee accommodation based on number of inhabitants, degree of housing deterioration and remoteness (distance to amenities) and assess the association with health in multivariate analyses. Accommodations with a moderate occupation, lowest levels of deterioration, and a central urban location showed the best health outcomes in terms of subjective general health status, depression, and general anxiety disorder (GAD). Associations were strongest for GAD and weakest for depression. Our findings inform policymakers on layout and location of refugee collective accommodation centres.

## Introduction

Living conditions of refugees and asylum seekers in resettlement countries are considered as post-migration conditions that can generate stress (1). While common mental disorders are highly prevalent in collective accommodation centres (2), studies rarely investigate the impact of the accommodation itself. Living in large reception centres significantly predicted mental problems among unaccompanied minors in the Netherlands (3); adult residents in state-provided reception centres suffered from poorer mental health than residents in other accommodation types (4). Ajlan considered collective accommodation centres as “unhomely places” in which the lack of privacy and autonomy creates uncertainty and desperation potentially causing violence among residents (5). Further, the Covid-19 pandemic has illustrated the particular physical health risks of living in collective accommodation centres (6,7).

While there is substantial evidence indicating that the type of accommodation can impact residents’ health, it is less well understood which contextual factors or combination thereof are most important in this association. Germany offers an informative setting to examine this question because of the broad range of accommodation types even within one federal state. In the past years, the number of refugees and asylum seekers living in German collective accommodation centres has declined (8,9), but every fourth was still living in collective accommodation at the end of 2019 (8,9). These centres can be divided into initial reception centres (including arrival- and AnkER-centres^1^) with compulsory residence until refugee status is determined (but for a maximum of 18 months), and subsequent collective accommodation centres^2^. The size and layout of German collective accommodation centres differ in many ways. In the federal state of North Rhine-Westphalia, for example, capacities of state-mandated reception centres ranged from 160 to more than 1000 people in 2019 (10). In 2016, many residents were housed in re-dedicated buildings, such as former offices, schools or hotels (11), each entailing specific and often poor housing conditions. Around one third were situated in rural areas and around one quarter in non-residential industrial areas (12). Moreover, accommodation centres often were located in non-residential areas, far away from infrastructure, contributing to the social exclusion of the residents (5).

In this paper, we devise a typology of refugee accommodation which helps to examine potential context-related health differences, using a large German federal state as example. We then analyse possible associations between accommodation types and the health of their residents.

## Data and methods

### Data

We analysed health and accommodation data of 412 refugees and asylum seekers from up to 58 different accommodation facilities in the federal state of Baden-Württemberg in the south-west of Germany, collected in the RESPOND study (12). The study provides a rich set of accommodation variables such as accommodation type, occupation density, geographical location, and deterioration of housing. For the latter, RESPOND researchers developed and validated an instrument, the “Small-area Housing Environment Deterioration” (SHED) index assessing different domains of housing and its physical environment (13). The geographical location was specified at district level. A binary variable indicated population density; < 150 inhabitants per km^2^ was considered as rural, else urban, following the standard BBSR^3^ definition. The distances to shops, medical and administrative services by foot and with public transport was estimated using Google Maps (GM) routes. Besides accommodation variables, the RESPOND study has also assessed a range of (subjective) physical and mental health outcomes, including a screening of GAD, depression symptoms, and the subjective general health status (12). They were measured using the General Anxiety Disorder GAD-2 (14), the Patient Health Questionnaire PHQ-2 for depression (15) as well as a general health indicator drawn from the European Health Interview Survey (EHIS, (16).

### Statistical methods

We performed a cluster analysis with the RESPOND variables that contained information about the accommodation and its physical context, i.e., the number of inhabitants, remoteness (walking distance to shops, medical or administrative services), urbanity (urban / rural distinction), and housing decay. For the latter, the overall SHED score as well as the normalized and z-standardized single scores were included in the analysis to identify housing differences. After exploring different cluster algorithms and methods, we chose a hierarchal, agglomerative clustering algorithm using Euclidean Distance and Ward’s method. We identified the number of clusters by visually assessing dendrogram and scatterplot, before adding the clusters as separate variables to the data set, representing different accommodation types. After bivariate analyses between clusters and health outcomes, we performed logistic regression analyses to obtain associations between the identified accommodation clusters and binary measures of general health, GAD, and depression adjusted for potential confounders. The independent variables included age, gender, and the cluster variables. In subsequent analyses, we assessed how the distribution of GAD and depression differed between the clusters. We created three distribution variables (GAD or depression present, both present, both absent) and used these distribution variables as dependent variables in additional logistic regression analyses. All statistical analyses were performed with R (version 3.6.2.).

## Results

### Clustering

Complete accommodation data of 411 refugees and asylum seekers were included in the cluster analysis. The resulting dendrogram and scatterplot pointed to either two or four clusters. To keep the within-group variance of the agglomerated clusters as high as possible, we decided to construct four clusters. A two-cluster solution would have led to a high level of dissimilarity. Cluster 1 and 3 were the smallest, containing 39 respondents of the RESPOND study each (from two and nine different accommodation centres, respectively). Cluster 4 contained 143 respondents from 29 different accommodation centres, and Cluster 2 190 respondents from 29 accommodation centres.

### Cluster characteristics

#### Cluster 1

Respondents in Cluster 1 had a mean age of 31.5 years (SD = 12), and 48.6% were female. In all other clusters, the proportion of women was lower. With an average of 151 residents per accommodation (SD = 9.0), Cluster 1 included the largest type of accommodation analysed here. The total SHED-score was 3.6 (SD = 1.5) which was higher than in all other clusters, indicating the highest level of housing deterioration. Besides the broken-windows indicator, all other single indicators scored equal to or larger than 0.5. Garbage accumulation and the outward appearance were most striking with a score of 0.8 each (SD = 0.2). Both accommodation centres in Cluster 1 were in an urban district according to the BBSR definition. Shops, medical and administrative services could all be reached by foot in around 15 minutes (SD ≤ 7.5).

Key characteristics of cluster 1:

- Largest accommodation type
- Highest state of decay
- Urban and central location

#### Cluster 2

Respondents in Cluster 2 had a mean age of 31.8 years (SD = 12). With a percentage of 22.5%, the proportion of women was the lowest of all clusters. This cluster had an average number of 70 residents per accommodation (SD = 20.9), including the second largest accommodation type. Housing deterioration was lower than in Cluster 1 but with a SHED-score of 1.1 (SD = 0.8) it was the cluster with the second highest state of decay. Garbage accumulation and the overall evaluation of the physical environment scored highest but with only 0.3 each (SD ≤ 0.3). Most of the 14 accommodation centres in this cluster were located in urban districts. Still, the percentage of residents living in urban areas was the lowest with only 72.1%. Shops and medical services could be accessed within approximately 10 minutes each by foot (SD ≈ 5), distance to administrative services was larger with around 22 minutes (SD = 10.3).

Key characteristics cluster 2:

- Second largest accommodation type
- Second highest state of decay
- Partly located in rural districts but still central

#### Cluster 3

In Cluster 3, respondents were slightly older than in the other clusters with a mean age of 33.1 years (SD = 10.4), 38.2% being female. With 15 residents on average (SD = 7.5), Cluster 3 presented the smallest accommodation type. It also showed one of the lowest SHED-scores with 0.8 (SD = 0.7). The overall evaluation of the physical environment scored highest at 0.3 (SD = 0.3), all other dimensions were close to zero. Though up to 90% of the accommodation centres (n = 9) can be considered as being located in urban districts, remoteness levels were remarkably high, with walking distances to shops, medical and administrative services being larger than 50 minutes (SD ≤ 15.3).

Key characteristics of cluster 3:

- Smallest accommodation type
- Low state of decay
- Located in urban districts but in the outskirts

#### Cluster 4

In Cluster 4, respondents were the oldest with a mean age of 33.4 years (SD = 9.9); 36.2% were female. This cluster included some of the smaller accommodation types with an average of 27 residents (SD = 13.6). With an overall SHED-score of 0.8 (SD = 0.9) and the highest single indicator being the overall evaluation of the physical environment (0.3, SD = 0.3), Cluster 4 presented quite similar low scores for housing deterioration as Cluster 3. The proportion of accommodation centres being located in urban districts was similarly low as in Cluster 2 with approximately 73%. Shops could be reached by foot within 11 minutes (SD = 8.3), medical services within 15 minutes (SD = 13.8) and administrative services within 17 minutes, though a standard deviation of 19.5 indicated relatively high variations here.

Key characteristics of cluster 4:

- Small accommodation type
- Low state of decay
- Partly located in rural districts but still central

More details of the cluster characteristics are provided in Table 1. In sum, the four clusters can roughly be divided into two larger accommodation types with deteriorated conditions and two smaller accommodation types with a low level of housing deterioration. The highest SHED-score was 3.6 out of a maximum of 6, indicating no extreme levels of housing deterioration. Scores for garbage accumulation were higher in the two clusters with a larger number of inhabitants, the same also applies to a lesser extent to the graffiti-score. In contrast, graffities and garbage accumulation were of little concern in the smallest accommodation cluster. All accommodation types were predominantly located in urban districts based on the urban-rural distinction applied here. Considering the level of remoteness, the results of Clusters 1, 2 and 4 were fairly similar, all showing acceptable distances to shops and essential services. Cluster 3, by contrast, was comparatively remote with walking distances beyond 50 minutes.

**Table 1:**
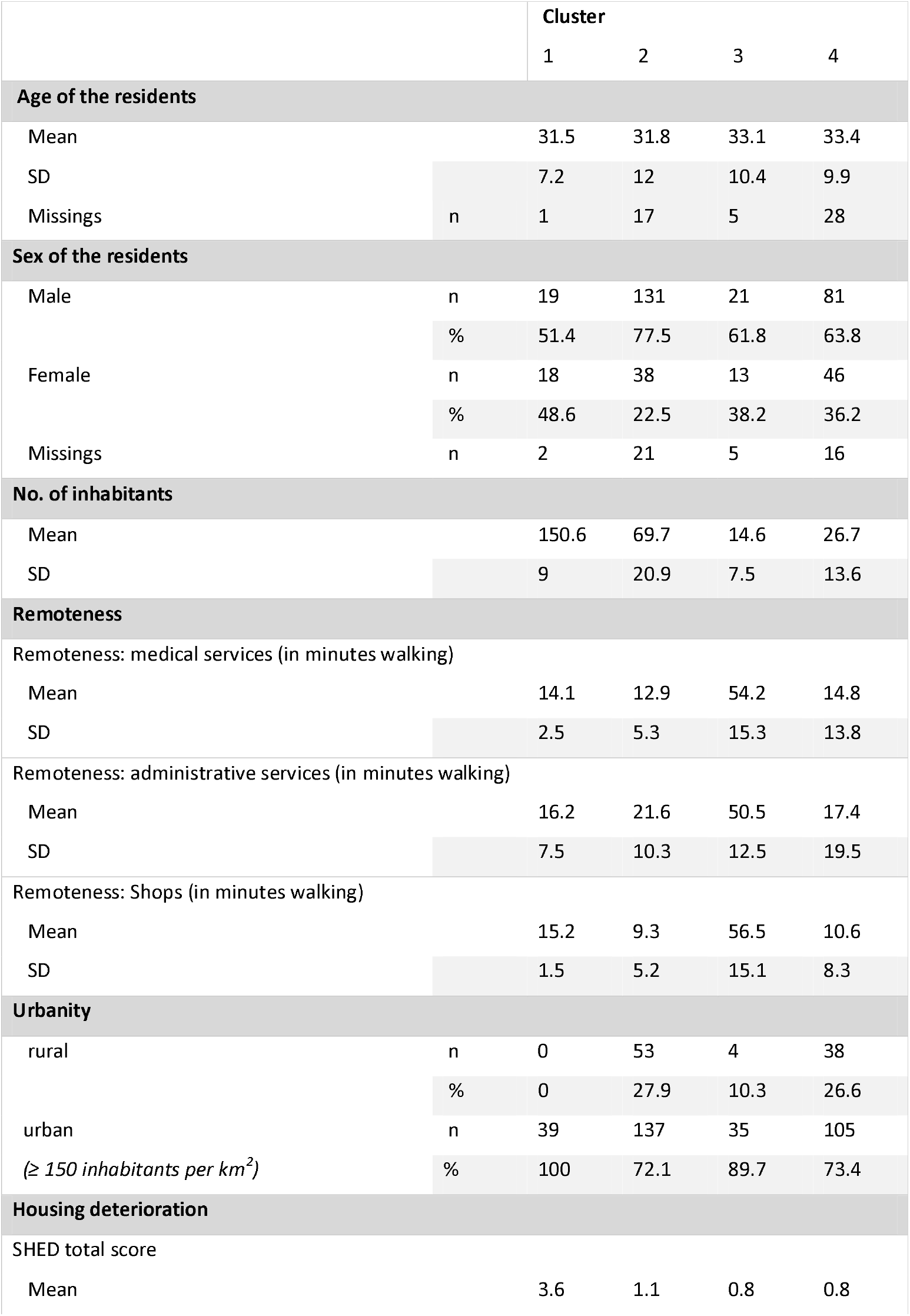
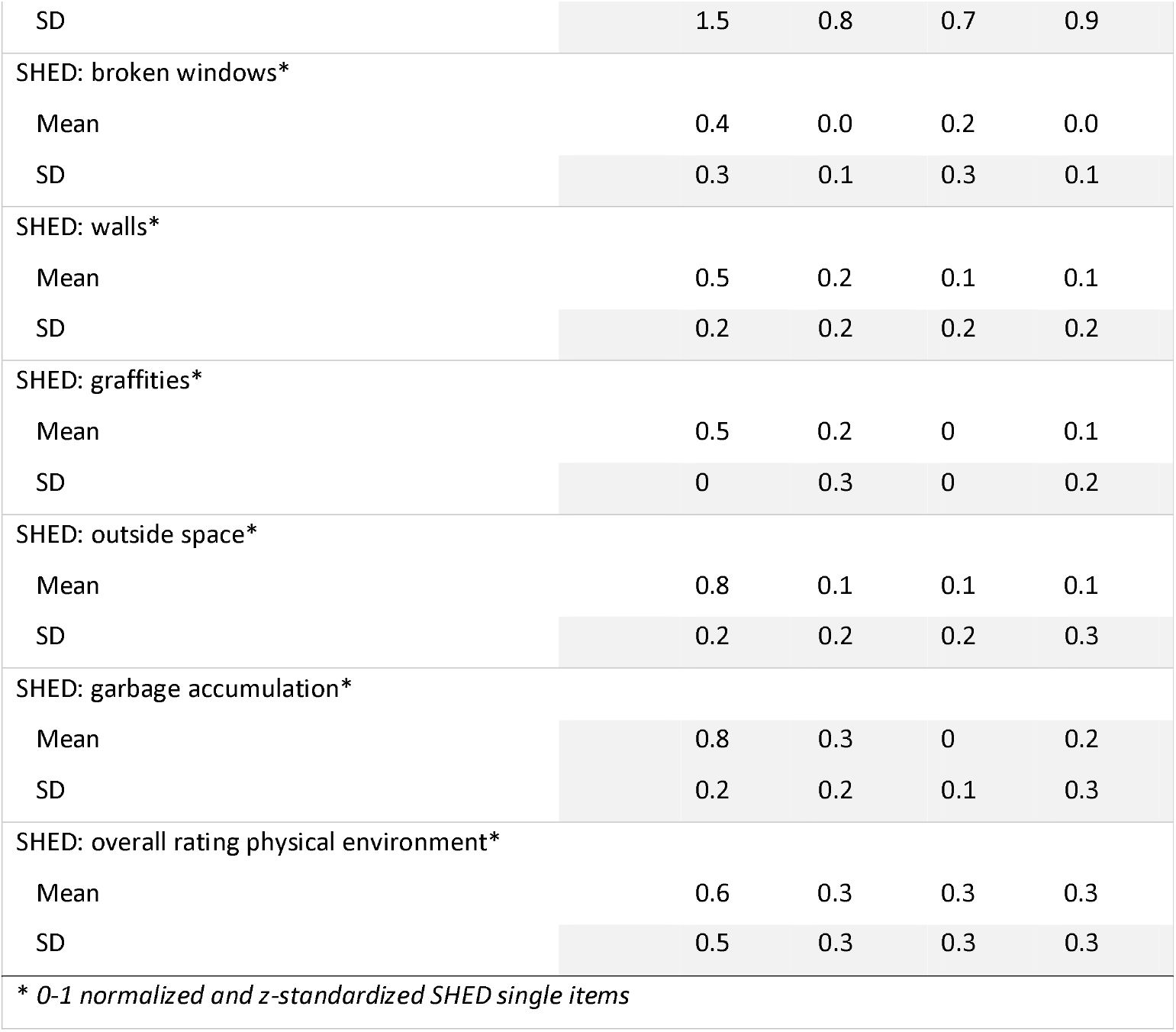
Cluster characteristics of refugee accommodation. Data: RESPOND study (11)

### Health disparities between clusters

The four clusters showed differences in the health status of their respondents. In all three health variables, residents in Cluster 4 showed better health outcomes than in the other clusters, as Table 2 illustrates. For the general subjective health status, only 12.3% of Cluster 4 residents indicated poor general health, while Cluster 3 scored the highest proportion with 25.7%. The differences were not statistically significant, though. Symptoms of impaired mental health were more common than of impaired general health in all clusters. In terms of GAD, more than one in two residents of Cluster 2 indicated symptoms. For Clusters 1 and 3, proportions were also relatively high, with more than 42%. Cluster 4 residents scored lower, around one third indicated symptoms here. The unadjusted differences in the frequencies of GAD symptoms between the clusters were statistically significant (χ^2^_(df = 3)_ = 13; *p* =0.005). The proportion of respondents with depression symptoms was lowest in Cluster 4 as well, with approximately 39%. In Clusters 1, 2 and 3, in contrast, around 1 in 2 people indicated symptoms of depression. Differences between the clusters were not statistically significant.

**Table 2:** Health characteristics of respondents grouped by accommodation cluster. Data: RESPOND study (11)

|  |  | Cluster |  |  |  |
| --- | --- | --- | --- | --- | --- |
|  |  | 1 | 2 | 3 | 4 |
| **General subjective health status** |  |  |  |  |  |
| Poor health | n | 6 | 38 | 9 | 16 |
|  | % | 16.7 | 22.5 | 25.7 | 12.3 |
| Missings | n | 3 | 21 | 4 | 13 |
|  | % | 7.69 | 11.05 | 10.26 | 9.09 |
| **General anxiety disorder (GAD2)** |  |  |  |  |  |
| GAD symptoms* | n | 14 | 86 | 18 | 38 |
|  | % | 42.4 | 53.8 | 48.6 | 32.2 |
| Missings | n | 6 | 30 | 2 | 25 |
|  | % | 15.38 | 15.79 | 5.13 | 17.48 |
| **Depression (PHQ-2)** |  |  |  |  |  |
| Depression symptoms | n | 17 | 79 | 17 | 45 |
|  | % | 51.5 | 49.1 | 47.2 | 38.5 |
| Missings | n | 6 | 29 | 3 | 26 |
|  | % | 15.38 | 15.26 | 7.69 | 18.18 |

\* differences statistically significant ( $p < 0.05$ )

### Results of regression analyses

An overview of the results of the regression analyses for poor general health, GAD, and depression (PHQ-2) is provided in Table 3. In the age- and gender-adjusted Model 1, Cluster 1, 2 and 3 all showed increased odds for residents having a poor general health status than Cluster 4. Cluster 2 showed the highest odd ratios here and was nearly significant at the 0.05-significance level (OR = 2.05; 95%-CI [1.00 – 4.46], p = 0.058). The adjusted Model 2 showed a significant association of GAD with accommodation type in Cluster 2 (OR = 2.7; 95%-CI [1.55 – 4.79], p = 0.001) and Cluster 3 (OR = 2.4; 95%-CI [1.04 – 5.64], p = 0.042) relative to Cluster 4. In the adjusted model for depression (Model 3), all clusters showed odd ratios larger than 1 relative to Cluster 4 but no association was statistically significant (p-values between 0.13 and 0.62).

**Table 3:** Accommodation cluster and health: Logistic regression results. Data: RESPOND study (11)

|  | Model 1: General health |  | Model 2: GAD |  | Model 3: PHQ-2 (Depression) |  |
| --- | --- | --- | --- | --- | --- | --- |
|  | OR (95% CI) | p-value | OR (95% CI) | p-value | OR (95% CI) | p-value |
| Constant | 0.08***<br>(0.03 - 0.23) | 0.000 | 0.37*<br>(0.15 - 0.88) | 0.026 | 0.35*<br>(0.15 - 0.81) | 0.016 |
| Age | 1.01<br>(0.99 - 1.04) | 0.313 | 1.00<br>(0.98 - 1.03) | 0.783 | 1.02<br>(1.00 - 1.04) | 0.081 |
| Gender | 1.51<br>(0.79 - 2.86) | 0.204 | 1.43<br>(0.85 - 2.42) | 0.174 | 1.07<br>(0.64 - 1.78) | 0.796 |
| Cluster 1 | 1.41<br>(0.41 - 4.28) | 0.556 | 1.25<br>(0.51 - 2.97) | 0.619 | 1.24<br>(0.53 - 2.89) | 0.620 |
| Cluster 2 | 2.05<br>(1.00 - 4.46) | 0.058 | 2.70***<br>(1.55 - 4.79) | 0.001 | 1.52<br>(0.88 - 2.63) | 0.134 |
| Cluster 3 | 1.69<br>(0.54 - 4.89) | 0.343 | 2.40*<br>(1.04 - 5.64) | 0.042 | 1.28<br>(0.55 - 2.96) | 0.558 |
| Nagelkerke $R^2$ | 0.035 | | 0.069* | | 0.025 | |
| AIC: | 275.9 |  | 387.2 |  | 397.4 |  |

Notes:

|  | Model 4: GAD + PHQ-2 = 1 |  | Model 5: GAD + PHQ-2 = 0 |  | Model 6: GAD OR PHQ-2 = 1 |  |
| --- | --- | --- | --- | --- | --- | --- |
|  | OR (95% CI) | p-value | OR (95% CI) | p-value | OR (95% CI) | p-value |
| Constant | 0.15***<br>(0.06 - 0.37) | 0.000 | 1.31<br>(0.56 - 3.13) | 0.530 | 0.64<br>(0.15 - 0.81) | 0.380 |
| Age | 1.02<br>(1.00 – 1.04) | 0.098 | 1.00<br>(0.97 - 1.02) | 0.650 | 0.98<br>(1.00 - 1.04) | 0.180 |
| Gender | 1.44<br>(0.84 - 2.47) | 0.188 | 0.91<br>(0.54 - 1.52) | 0.712 | 0.73<br>(0.64 - 1.78) | 0.320 |
| Cluster 1 | 0.99<br>(0.35 – 2.57) | 0.988 | 0.66<br>(0.29 - 1.53) | 0.334 | 1.68<br>(0.53 - 2.89) | 0.260 |
| Cluster 2 | 2.23**<br>(1.24 – 4.10) | 0.009 | 0.50*<br>(0.29 - 0.86) | 0.013 | 0.99<br>(0.88 - 2.63) | 0.970 |
| Cluster 3 | 2.01<br>(0.85 – 4.95) | 0.107 | 0.62<br>(0.27 - 1.43) | 0.270 | 0.80<br>(0.55 - 2.96) | 0.670 |
| *Nagelkerke R<sup>2</sup>* | 0.062* |  | 0.031 |  | 0.026 |  |
| *AIC:* | 360.3 |  | 390 |  | 323.6 |  |

### Distribution of depression and GAD in the clusters

In the sample, depression and anxiety disorders were correlated (r = 0.52). We additionally assessed the distribution of depression and GAD in the clusters (see Figure 1) and analysed how regression results changed when using these distribution variables as dependent variables (see Table 3). Higher proportions of residents in all clusters showed both symptoms than GAD symptoms without depression, or vice versa. Only cluster differences found for both symptoms present were statistically significant (χ^2^_(df = 3)_ = 8; *p* =0.05).

**Figure 1:**
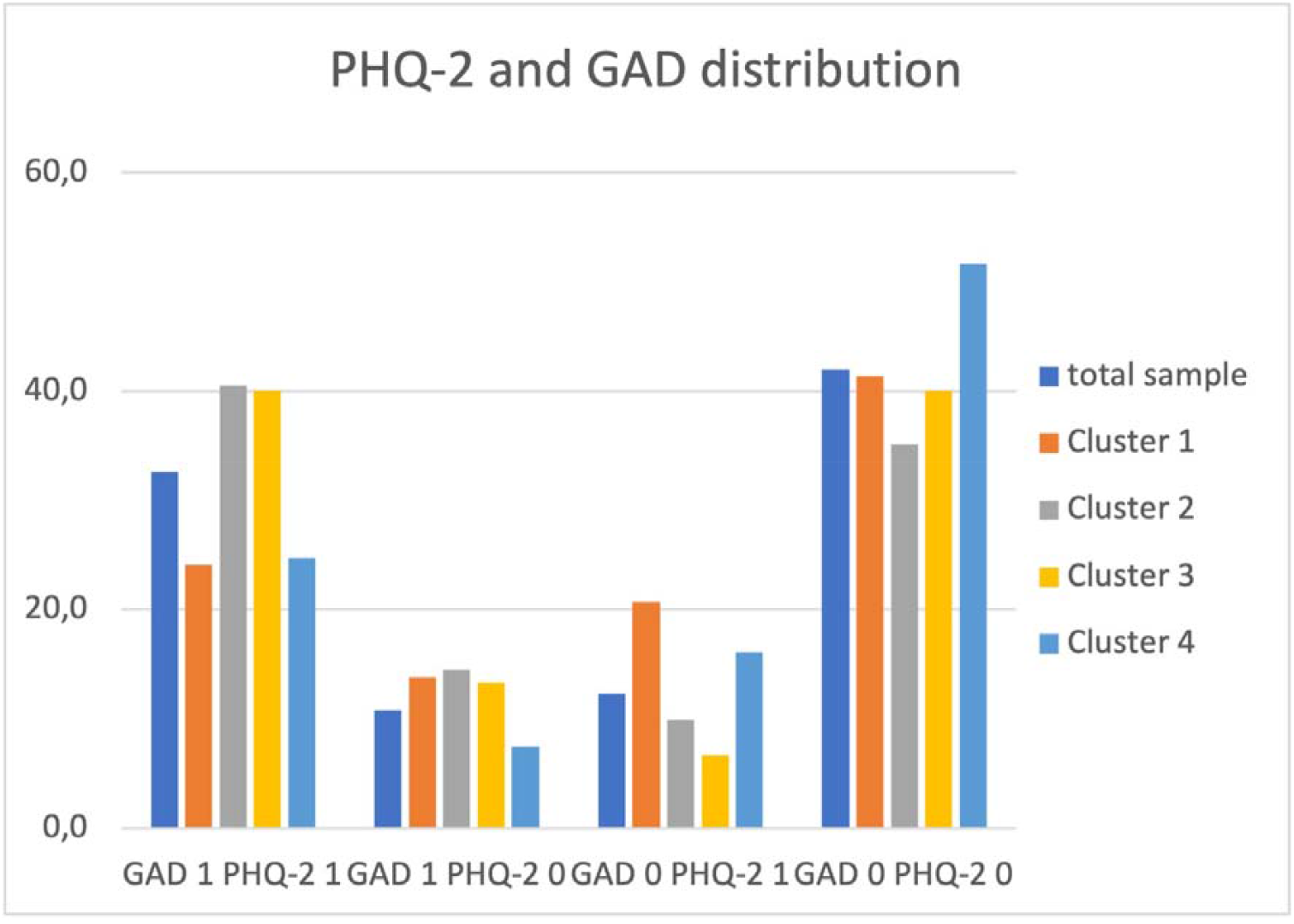
Distribution of depression (PHQ-2) and GAD symptoms in the total sample and grouped by cluster. Data: RESPOND study (11)

In the regression analyses, we found age- and gender-adjusted significant differences between Cluster 2 and 4 (OR = 2.3; 95%-CI [1.24 – 4.10]; p = 0.009) in the model for comorbid GAD and depression (Model 4, Table 3). A model for the absence of both conditions (Model 5) supported these results, again indicating significant effects for Cluster 2. Further, a model for the presence of either GAD or depression symptoms could not identify any statistically significant cluster effects.

## Discussion

Collective refugee accommodation centres are considered to be postmigration stress factors impacting health (17). However, to the best of our knowledge, empirical research investigating health impacts of collective accommodation has put little emphasis on contextual differences in the accommodation setting as such.

In Germany, collective accommodation is subject to substantial variability regarding size, location, and the quality of the housing environment. We aimed to devise a typology of refugee accommodation by conducting a cluster analysis and to analyse associations between different accommodation types and health. We identified four accommodation clusters: 1) a large accommodation type, located in central urban areas with visible deteriorated housing conditions (mean total SHED = 3.6/6); 2) a large-to-medium sized accommodation type frequently located in rural areas (though still located close to amenities) with few deteriorated conditions (mean total SHED = 1.1 /6); 3) a small accommodation type, predominantly located in urban areas with negligible levels of housing deterioration (mean total SHED = 0.8 /6) but remotely located from important amenities; and 4) a small to medium sized accommodation type located partially in rural areas but close to amenities and with negligible levels of housing deterioration (mean total SHED = 0.8 /6).

Our analyses consistently show that the best health outcomes could be found where residents lived in less crowded, less deteriorated, and less remotely located accommodation centres. There, residents suffer less frequently from GAD and depression, and more often report to be in good general health. In subsequent analyses, we also found that overall, a high proportion of residents showed comorbid symptoms of GAD and depression, odds for unfavourable health outcomes being significantly higher in Cluster 2 compared to Cluster 4. This is consistent with other findings pointing to comorbidity of depression and anxiety disorder (18–20).

Assuming the contextual differences in Cluster 2 and 3 compared to Cluster 4 are associated with the health outcomes in the clusters, issues of crowded dwellings, housing deterioration but also remoteness should be investigated more closely. Studies assessing the health impact of housing among refugee and asylum seeker populations in resettlement countries frequently point to issues of overcrowding, especially when the accommodation must be shared with strangers and health needs vary between the residents (21). Evans et al. (22) found a consistent positive relationship between overcrowding (measured by number of rooms per people) and psychological distress in different institutionalized populations (e.g. among prisoners or college students). The authors also pointed to indirect pathways linking the built environment to mental problems. More crowded residential settings can lead to social withdrawal with negative consequences for the development and maintenance of socially supportive relationships.

Research investigating health impacts of the physical environment of refugee accommodation is yet sparse, but our findings are supported by evidence from the literature assessing neighbourhood disorder. A meta-analysis by O’Brien et al. (23) found a persistent association of neighbourhood disorder with unfavourable mental health outcomes as well as overall health. The results support the psychosocial model of disadvantage, indicating that stressful conditions – such as deteriorated neighbourhoods - can affect mental health, though the studies assessed were mainly cross-sectional, thus not allowing to confirm causality. We also found that among the different aspects of housing deterioration, garbage accumulation showed a strong effect. More research on this association is needed (24).

We did not find consistent evidence whether and how remoteness, i.e. the distance to important amenities, is associated with health. While an association between urban environment and poorer mental health outcomes is well documented, there is low evidence that remoteness is associated with mental illness. Peen et al. (25) argued that a range of stress factors potentially harmful for mental health are more prevalent in urban areas than in rural areas, i.e. social factors such as life events and social isolation, or physical factors such as crowded housing, high population density, and air pollution. Adli and Schöndorf (26) support the assumption that urban districts induce social stress which in turn can increase the risk of mental illness. While Eckert et al. (27) and Goldney et al. (28) found no associations between remoteness and mental illness in Australia, Kelly et al. (29) found higher risk of mental problems for residents living in more remote areas. In all three studies, remoteness was defined by road distance to services. Kelly et al. (29) pointed to the geographic variabilities in mental health but also raised the question whether the observed association may be less due to the remoteness itself but rather due to associated regional characteristics such as social isolation, community size, or socioeconomic position.

### Strengths and limitations

We have addressed contextual variabilities of collective refugee accommodation centres in Germany to investigate associations between housing and refugee health in a comparative way. By using a cluster analysis approach, we identified four distinct types of refugee accommodation. This enabled us to go beyond regular multivariate analyses based on individual contextual characteristics and focus on contextual combination of different characteristics when assessing their associations with health and presenting health disparities.

Cluster size varied considerably, ranging from 39 cases to 190 respondents. Since one of the smaller clusters was consistent and repeatedly occurred in different pre-analyses with varying clustering approaches, we decided to accept the variability in cluster size (and the small number of accommodation centres cluster 1 was based on), being aware that the clusters can be compared with each other only to a limited extent.

In regression analyses, only few effects were found to be statistically significant. This may be due to the small sample sizes in some of the clusters. Especially Cluster 1 failed to yield significant associations relative to Cluster 4. Another statistical issue was missing values in the data for GAD and depression. Whereas the percentages of missings were relatively even in Cluster 1, 2 and 4, Cluster 3 had lower rates of missings (see Table 2). Since Cluster 4 contained the highest number of missings overall with 17.5% (GAD) and 18.3% (depression), associations found in this analysis could have been weaker and frequencies of residents showing GAD and depression symptoms could have been higher.

Further, we could identify few cluster differences regarding urbanity. Given our definition of urbanity, it is rather unsurprising that each accommodation type identified here was predominantly located in urban districts. For example, in Baden Württemberg, where data has been collected, overall population density was 311 inhabitants per km^2^ in 2019 (30), far above the cut-off applied here. However, as refugees are distributed to districts according (among other factors) their population size, the majority of refugees will be accommodated in more urban areas.

Also, we could not investigate to which extent the residents of the larger accommodation types lived in overcrowded settings according to the Eurostat definition since the data did not provide information about room numbers of the accommodation centres assessed. However, with an average number of residents of up to 150, we can assume that the specifications of the Eurostat definition are not met, and that residents of these accommodations (clusters 1 and 2) share rooms. As the Eurostat definition applies to private households rather than to collective accommodation centres, a more appropriate concept should be developed.

The cross-sectional design of the data allowed us to identify only associations between the accommodation cluster and unfavourable health outcomes. Longitudinal approaches are needed to assess whether the accommodation types are causally associated with certain health outcomes. O’Brien et al. (23) for example argued that poor mental health may also result in more negative assessments of the physical environment. Since the SHED index was assessed by researchers, not by residents themselves, this form of same-source bias is unlikely here. It may be possible, though, that researchers would consider the accommodation as run-down while refugees might appreciate the conditions compared to other accommodation settings they have experienced during flight.

Future research should also account for the stage of the asylum application process of interviewees. For example, it is possible that Cluster 4 contained a lower proportion of residents whose asylum claim was still under process, which has been found to be a significant predictor of mental health problems (31). Since asylum seekers live more often in deprived areas with unfavourable living conditions (32) like a run-down physical environment, the pathway of how exactly accommodation type impacts health warrants further research.

## Conclusion

The context in which people live can affect their health in many ways. By conducting a cluster analysis of variables related to refugee accommodation in Germany, we have identified four different accommodation types reflecting contextual differences in the number of inhabitants, the degree of housing deterioration, and the distance to important amenities, thereby contributing to the development of a typology of refugee accommodation. The accommodation type with a moderate number of occupants, lowest levels of deterioration, and a central and urban location was associated with favourable health outcomes in terms of subjective general health status, depression, and GAD – the latter most strongly. The odds of unfavourable health increased for accommodation clusters that either showed a high state of decay and a large accommodation size (Cluster 2), or a small accommodation size remote from important amenities (Cluster 3). Effects were weakest for depression. Subsequent analyses showed, though, that when assessing comorbidity of GAD and depression, the larger, more deteriorated accommodation type (Cluster 2) yielded similar significant effects compared to the analysis of GAD alone. The findings from this study should be considered in political decisions on the layout as well as the choice of location of collective accommodation centres in Germany and comparable countries.

## Data Availability

Data are available from the authors upon reasonable request.

https://respond-study.org/en/resources/

## Footnotes

1 AnkER - centres are special types of reception centres concentrating all relevant actors for the reception, distribution across Germany and (where relevant) the deportation of asylum seekers in one site. This concept is mainly implemented in Bavaria.

2 In the following the term “collective accommodation centre” is used to denote initial reception centres as well as subsequent collective accommodation unless a distinction is necessary.

3 Federal Institute for Research on Building, Urban Affairs and Spatial Development (BBSR)

